# Assessment of clinical characteristics and mortality-associated factors in COVID-19 Critical cases in Kuwait

**DOI:** 10.1101/2020.06.17.20134007

**Authors:** Mariam Ayed, Abdulwahab A Borahmah, Anwar Yazdani, Ahmad Sultan, Ahmad Mossad, Hanouf Rawdhan

## Abstract

**Purpose:** To assess the clinical characteristics and identify mortality predicting factors in intensive care unit (ICU)-admitted COVID-19 confirmed patients.

**Methods:** We recruited and analyzed COVID-19 infected adult patients (age≥ 18 years) who were admitted to the ICU at Jaber AlAhmad Al Sabah Hospital,Kuwait, between 1^st^March, 2020and 30^th^April 2020. The patients were followed up to 20^th^ May, 2020. The risk factors associated with in-hospital mortality were assessed using multiple regression analysis.

**Results:** We recruited a total of 103 ICU patients in this retrospective cohort. The median age of the patients was 53 years (Interquartile Range (IQR): 44-63 years). The fatality rate was 43.7%. Among the patients, majority were males (85.5%) and 38% patients had more than two comorbidities. Pre-existing hypertension (OR:3.2,95%CI: 1.2-8.9), moderate/severe ARDS (OR: 3.4, 95%CI: 1.1-10.8),lymphocyte counts <0.5 (OR: 6.1, 95%CI: 1.2-29.8)albumin < 22 (OR: 7.5, 95%CI: 2.1-26.2), procalcitonin >0.2 (OR: 3.8, 95%CI: 1.3-7.8), D-Dimer >1200 (OR: 5.1, 95%CI: 1.2-21.6), and the need forcontinuous renal replacement therapy (OR: 9.3, 95%CI: 2.4-36.2) weresignificantly associated with mortality.

**Conclusion:** This study describes the clinical characteristics and predictors ofmortality among ICU patients. Early identification of risk factors to mortality might help in their better outcome.

## 1. Introduction

The outbreak of novel coronavirus disease began with a simple case of pneumonia in Wuhan, China, in late December, 2019. By 30^th^ January, 2020, World Health Organization (WHO) declared the disease as a Public Health Emergency of International Concern and renamed it as COVID-19 [1]. Less than a month later, on February 24^th^, the State of Kuwait confirmed 3 patients on a flight coming from Islamic Republic of Iran to have contracted the virus, marking the start of the outbreak in the country [2]. By 3^rd^ March, 2020, the first COVID-19 patient was admitted to the ICU. In the next couple of months, more than 100 COVID-19 confirmed cases got admitted to the ICU.

The COVID-19 pandemic is considered one of the greatest global public health crisis since the 1918 influenza pandemic [3]. Although the clinical spectrum of the infection appears to be broad, patients infected with COVID-19 are found to be at high risk of developing pneumonia, ARDS, or multi-organ failure that requires hospitalization and admission under a critical care unit [4]. The major etiology agent of COVID-19 is severe acute respiratory syndrome coronavirus 2 (SARS-CoV-2). Coronaviruses are minute single-stranded RNA viruses that belong to a broad family named Coronaviridae. Earlier, these viruses were believed to infect only animals. In 2002, the world saw the first instance of human coronavirus infection with the spread of severe acute respiratory syndrome (SARS) in China. The main etiological agent responsible for SARS was identified to be SARS-CoV [5]. The next major coronavirus-related disease was the Middle East Respiratory Syndrome (MERS) that arose in Saudi Arabia in 2012, which was caused by the MERS-CoV [6].The SARS-CoV-2 is closely related to the SARS-CoV; the difference between the two viruses lies in the mortality and transmission rates. The transmission rate of SARS-CoV-2 is much higher than that of SARS-CoV as well as MERS-CoV. However, SARS-CoV-2 has much lower mortality rate (∼2.9%) compared to that of SARS-CoV (9%).

However, even after encountering two coronavirus-related diseases, the lack of adequate preparedness and late implementation of the countermeasures led to a drastic global spread of COVID-19 within a short period of time. Furthermore, lack of an appropriate vaccine has also contributed to the spread of this disease and the high mortality, despite low mortality rate of the virus itself. Several epidemiological studies have shown that advanced age and presence and history of severe comorbidities, such as cardiovascular diseases, are significantly associated with COVID-19-related mortality. Moreover, COVID-19-related mortality has been attributed to virus-activated “cytokine storm syndrome” [7].

Till date, several studies are still being conducted to elucidate the epidemiology and prevalence of COVID-19 across different countries. The high rate of spread of this disease has put a lot of burden on an already overwhelmed healthcare system. This has led to development of new COVID-19-specific treatment centers and prioritizing the use of medical resources based on patient characteristics. Understanding the risk factors ofintensive care unit (ICU) admission is an important factor in triage decision making and identification of the patients that might benefit more from critical care services [8]. Here, we assessed the clinical characteristics and identified the mortality-associated predictive factors in COVID-19 patients of Kuwait who were admitted to the ICU. To the best of our knowledge, there is no literature that describes the clinical characteristics of ICU-admitted COVID-19 patients from our demographic region.

### 2. Methods

#### 2.1. Study design and settings

We conducted a retrospective study consisting of patientsadmitted to the ICU at Jaber Al-Ahmad Hospital, between 1^st^March 2020 and 30^th^April 2020.The patient outcome was followed up till 20^th^May 2020. Jaber AlAhmad Al Sabah hospital was designated as national hospital for the management of COVID-19 infected patients in Kuwait. The hospital initially hadthree ICUs (120 beds) and, as the pandemic progressed, the ICU bed capacity was increased to 145 beds.

#### 2.2. Patient criteria

We enrolled patients aged ≥18 years admitted to ICU due to COVID-19 infection as confirmed by real-time reverse transcriptase polymerase chainreaction (RT-PCR) assay of nasopharyngeal swab specimens. Management and treatment of COVID-19 ICU patients were according to the published international and local guidelines [9].

#### 2.3. Data collection and definition

Patient data wereextracted from the electronic medical records using standardized data collection sheet. We recorded the information on pre-existing medical condition including hypertension, diabetes, ischemic heart disease, hyperlipidemia, bronchial asthma, chronic renal failure,malignancy, and autoimmune diseases), home medications, exposure history (i.e., travel or sick contact), and presenting symptoms prior to admission, such as fever, sore throat/ nasal or sinus congestion, cough/chest pain, vomiting/diarrhea, and fatigue/myalgia. We collected the laboratory and radiological test results upon admission to ICU. Chest x-ray and CT-scan findings were categorized as normal, unilateral infiltration, bilateral infiltration, and ground glass appearance.CT-scan chest and echocardiographs were requested as clinically indicated and at the discretion of the treating physicians. We also recorded data on maximum respiratory support (i.e.. low flow nasal cannula, non-invasive mechanical ventilation and intubation) and additional adjunctive support, including inhaled nitric oxide, inotropes, renal replacement therapy (RRT) and extracorporeal membrane oxygenation (ECMO). Information on patient-specific therapies, such as administration of hydroxychloroquine, antivirals, and convalescent plasma,was also collected.

Acute respiratory distress syndrome (ARDS) was defined according to the Berlin guidelines[10], and the quick Sequential Failure Assessment (qSOFA) score was calculated upon admission according to the Third International Consensus Definitions Task Force for sepsis and septic shock [11]. Secondary infection was defined as clinical signs of pneumonia or bacteremia and positive culture on blood or body fluid, or endotracheal or bronchoalveolar lavage specimens. The study protocol was approved by the Ethics Committee of the Ministry of Health of Kuwait (IRB 2020/1411).

#### 2.4. Statistical analysis

We reported continuous variables as median (interquartile range [IQR]) and categorical variables as number and percentage. With mortality as the outcome of interest, differences between the deceased patients and the discharged patients were analyzed using Fisher’s exact test for categorical variables and Mann-Whitney U test for continuous variables.

We dichotomized the laboratory findings based on the clinical assessment and normal references. We used multivariate logistic regression model to identify predictors of mortality. Variables that were found to be significant during the univariate analysis were included in the multivariate regression model. Age was included in the final model as it had been described to be a risk factor for mortality in previous studies.[7]Variables with more than 50% missing values were not included in the model (such as, high sensitivity cardiac troponin). Final model fitting was evaluated using Hosmer-Lemeshow goodness-of-fit test(P value=0.468). Statistical analysis was performed using STATA/IC 14 (STATA Corp, College Station, Texas), and 2-tailed *P* < 0.05 was considered statistically significant without adjustment for multiple testing.

## 3. Results

As of May 20^th^ 2020, among the 103 patients, 47 (45.6%)patients have been discharged, 45 (43.7%) patients died, and 11 (10.7%) patients remained in ICU.

### 3.1. Demographic and clinical characteristics

The median age was 53 years [IQR: 44-63] and majority of the patients (85.5% [88/103]) were male. Two or more comorbidities were present in 37% (38/103) of the patients with diabetes mellitus being the most common (in 39% (40/103) patients) followed by hypertension (35%(36/103) patients).About 33%, 12.7%, and 9.8% of the patients were on insulin, beta blockers, and angiotensin converting enzyme inhibitors (ACEI), respectively. The most common presenting symptoms were fever (61%), followed by cough/chest pain (48%). Travel history and contact exposure with a COVID-19 infected person were documented in 28% of the patients. The median duration of symptoms was 2 days [IQR:1-5] and 5 days [IQR:1-8] prior to hospital and ICU admission, respectively (Table 1).

**Table 1:**
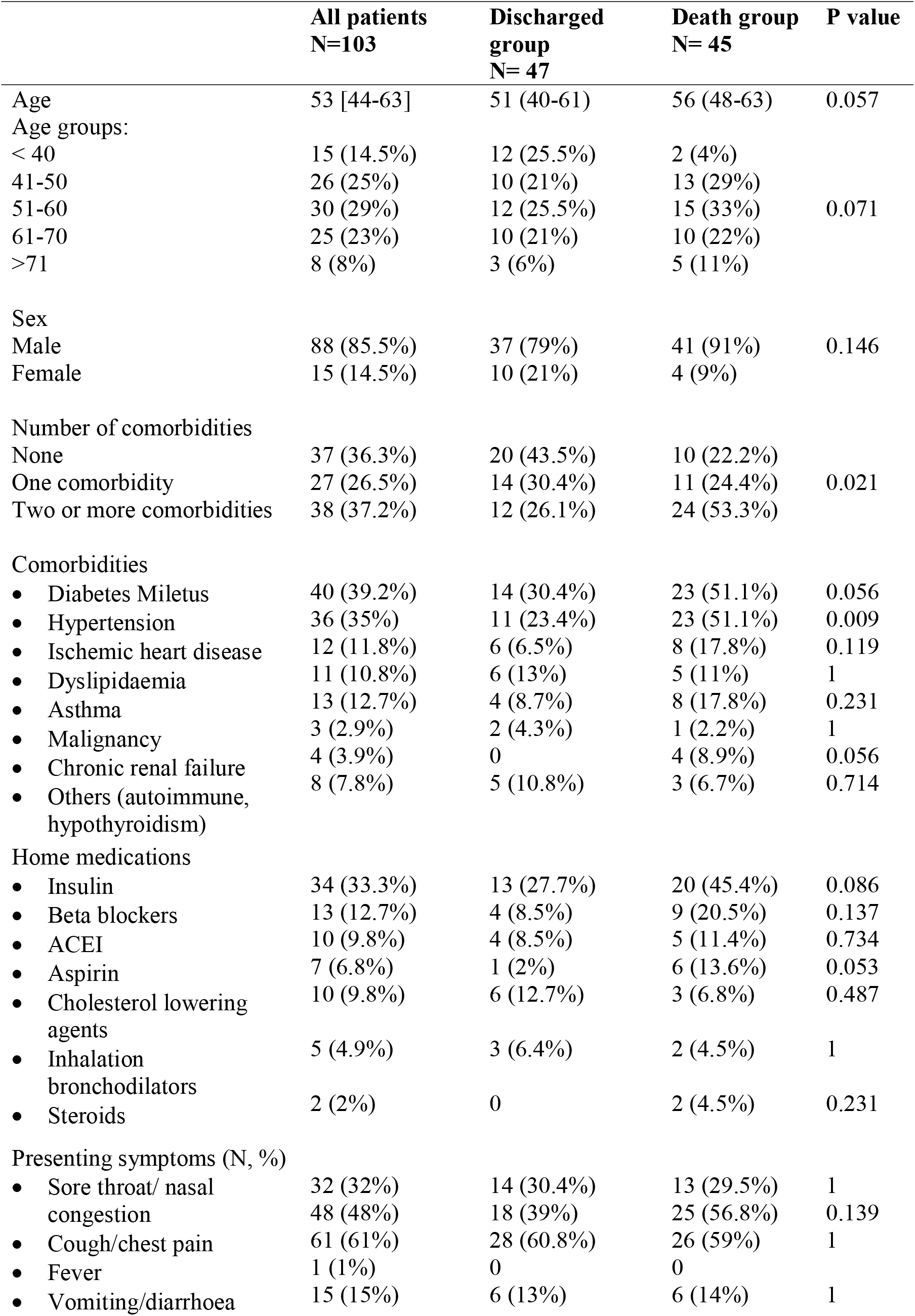
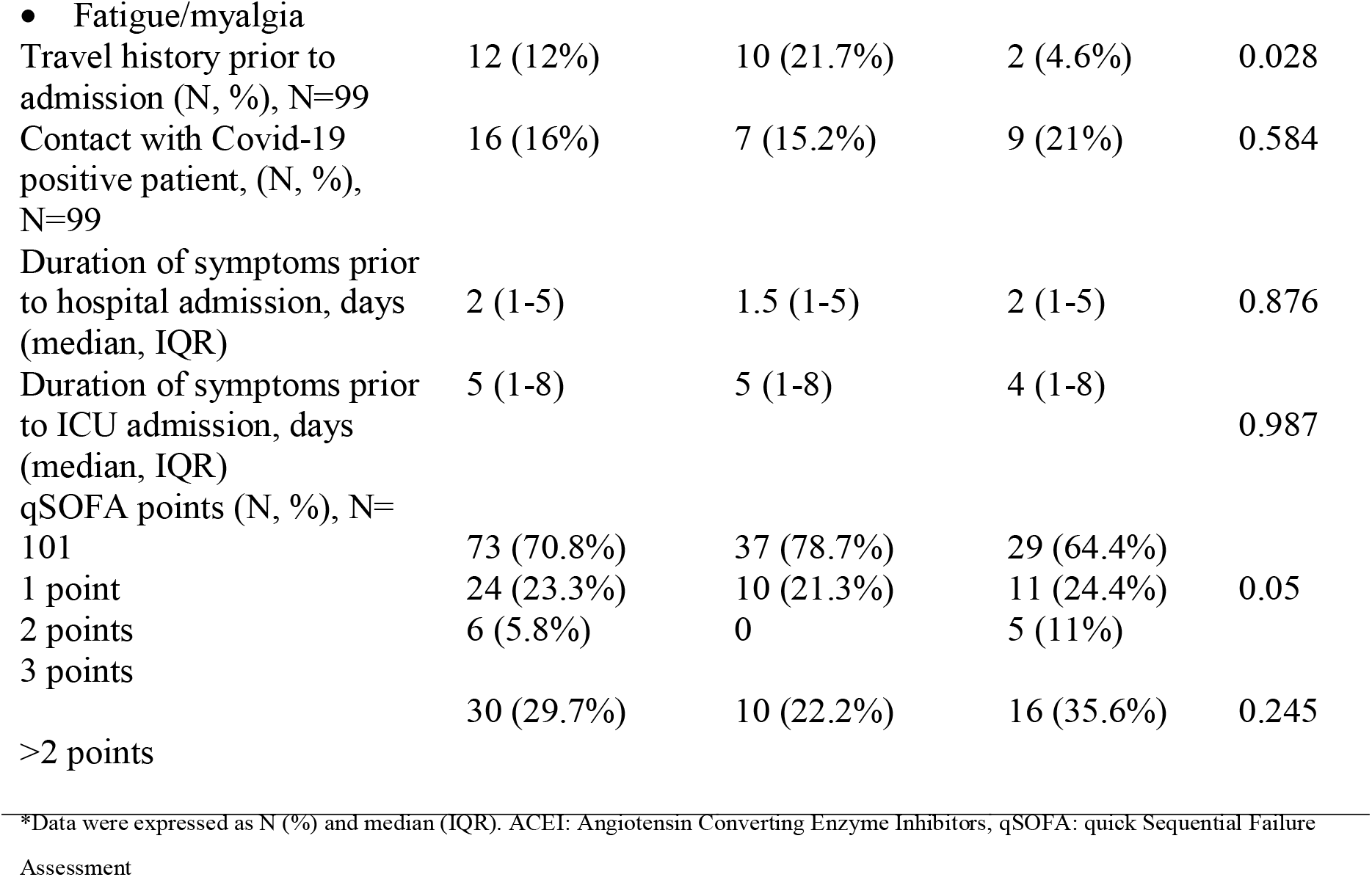
Baseline demographic and clinical characteristics.

|  | **All patients**<br>**N=103** | **Discharged**<br>**group**<br>**N= 47** | **Death group**<br>**N= 45** | **P value** |
| --- | --- | --- | --- | --- |
| Age | 53 [44-63] | 51 (40-61) | 56 (48-63) | 0.057 |
| Age groups: |  |  |  |  |
| < 40 | 15 (14.5%) | 12 (25.5%) | 2 (4%) |  |
| 41-50 | 26 (25%) | 10 (21%) | 13 (29%) |  |
| 51-60 | 30 (29%) | 12 (25.5%) | 15 (33%) | 0.071 |
| 61-70 | 25 (23%) | 10 (21%) | 10 (22%) |  |
| >71 | 8 (8%) | 3 (6%) | 5 (11%) |  |
| Sex |  |  |  |  |
| Male | 88 (85.5%) | 37 (79%) | 41 (91%) | 0.146 |
| Female | 15 (14.5%) | 10 (21%) | 4 (9%) |  |
| Number of comorbidities |  |  |  |  |
| None | 37 (36.3%) | 20 (43.5%) | 10 (22.2%) |  |
| One comorbidity | 27 (26.5%) | 14 (30.4%) | 11 (24.4%) | 0.021 |
| Two or more comorbidities | 38 (37.2%) | 12 (26.1%) | 24 (53.3%) |  |
| Comorbidities |  |  |  |  |
| • Diabetes Miletus | 40 (39.2%) | 14 (30.4%) | 23 (51.1%) | 0.056 |
| • Hypertension | 36 (35%) | 11 (23.4%) | 23 (51.1%) | 0.009 |
| • Ischemic heart disease | 12 (11.8%) | 6 (6.5%) | 8 (17.8%) | 0.119 |
| • Dyslipidaemia | 11 (10.8%) | 6 (13%) | 5 (11%) | 1 |
| • Asthma | 13 (12.7%) | 4 (8.7%) | 8 (17.8%) | 0.231 |
| • Malignancy | 3 (2.9%) | 2 (4.3%) | 1 (2.2%) | 1 |
| • Chronic renal failure | 4 (3.9%) | 0 | 4 (8.9%) | 0.056 |
| • Others (autoimmune, hypothyroidism) | 8 (7.8%) | 5 (10.8%) | 3 (6.7%) | 0.714 |
| Home medications |  |  |  |  |
| • Insulin | 34 (33.3%) | 13 (27.7%) | 20 (45.4%) | 0.086 |
| • Beta blockers | 13 (12.7%) | 4 (8.5%) | 9 (20.5%) | 0.137 |
| • ACEI | 10 (9.8%) | 4 (8.5%) | 5 (11.4%) | 0.734 |
| • Aspirin | 7 (6.8%) | 1 (2%) | 6 (13.6%) | 0.053 |
| • Cholesterol lowering agents | 10 (9.8%) | 6 (12.7%) | 3 (6.8%) | 0.487 |
| • Inhalation bronchodilators | 5 (4.9%) | 3 (6.4%) | 2 (4.5%) | 1 |
| • Steroids | 2 (2%) | 0 | 2 (4.5%) | 0.231 |
| Presenting symptoms (N, %) |  |  |  |  |
| • Sore throat/ nasal congestion | 32 (32%) | 14 (30.4%) | 13 (29.5%) | 1 |
|  | 48 (48%) | 18 (39%) | 25 (56.8%) | 0.139 |
| • Cough/chest pain | 61 (61%) | 28 (60.8%) | 26 (59%) | 1 |
| • Fever | 1 (1%) | 0 | 0 |  |
| • Vomiting/diarrhoea | 15 (15%) | 6 (13%) | 6 (14%) | 1 |
| • Fatigue/myalgia |  |  |  |  |
| Travel history prior to admission (N, %), N=99 | 12 (12%) | 10 (21.7%) | 2 (4.6%) | 0.028 |
| Contact with Covid-19 positive patient, (N, %), N=99 | 16 (16%) | 7 (15.2%) | 9 (21%) | 0.584 |
| Duration of symptoms prior to hospital admission, days (median, IQR) | 2 (1-5) | 1.5 (1-5) | 2 (1-5) | 0.876 |
| Duration of symptoms prior to ICU admission, days (median, IQR) | 5 (1-8) | 5 (1-8) | 4 (1-8) | 0.987 |
| qSOFA points (N, %), N=101 | 73 (70.8%) | 37 (78.7%) | 29 (64.4%) |  |
| 1 point | 24 (23.3%) | 10 (21.3%) | 11 (24.4%) | 0.05 |
| 2 points | 6 (5.8%) | 0 | 5 (11%) |  |
| 3 points | 30 (29.7%) | 10 (22.2%) | 16 (35.6%) | 0.245 |
| >2 points |  |  |  |  |

\*Data were expressed as N (%) and median (IQR). ACEI: Angiotensin Converting Enzyme Inhibitors, qSOFA: quick Sequential Failure Assessment

There was no significant difference in age, gender, and nationality between the discharged and deceased patients. Deceased patients were more likely to have more than two comorbidities compared to the discharged patients (56% versus 27%; P = 0.017). Higher proportion of deceased patients were hypertensive compared to discharged group (56% versus 24%; P =0.003). Travel history was less likely to be reported among the deceased patients compared to the discharged patients (4.6% versus 21.7%; P =0.028). Moreover, deceased patients tended to exhibit higher qSOFA score than the discharged patients (P =0.05) (Table 1).

### 3.2. Laboratory and radiological findings

Table 2 shows the laboratory and radiological findings of the patients on admission to ICU. Total lymphocyte countsand serum albumin levels were significantly lower among the deceased patients. Levels of creatinine, procalcitonin (PCT), C-reactive protein CRP), D-dimer, and high sensitivity (HS)cardiac troponin were significantly elevated in the deceased patients compared to the discharged patients.

**Table 2:**
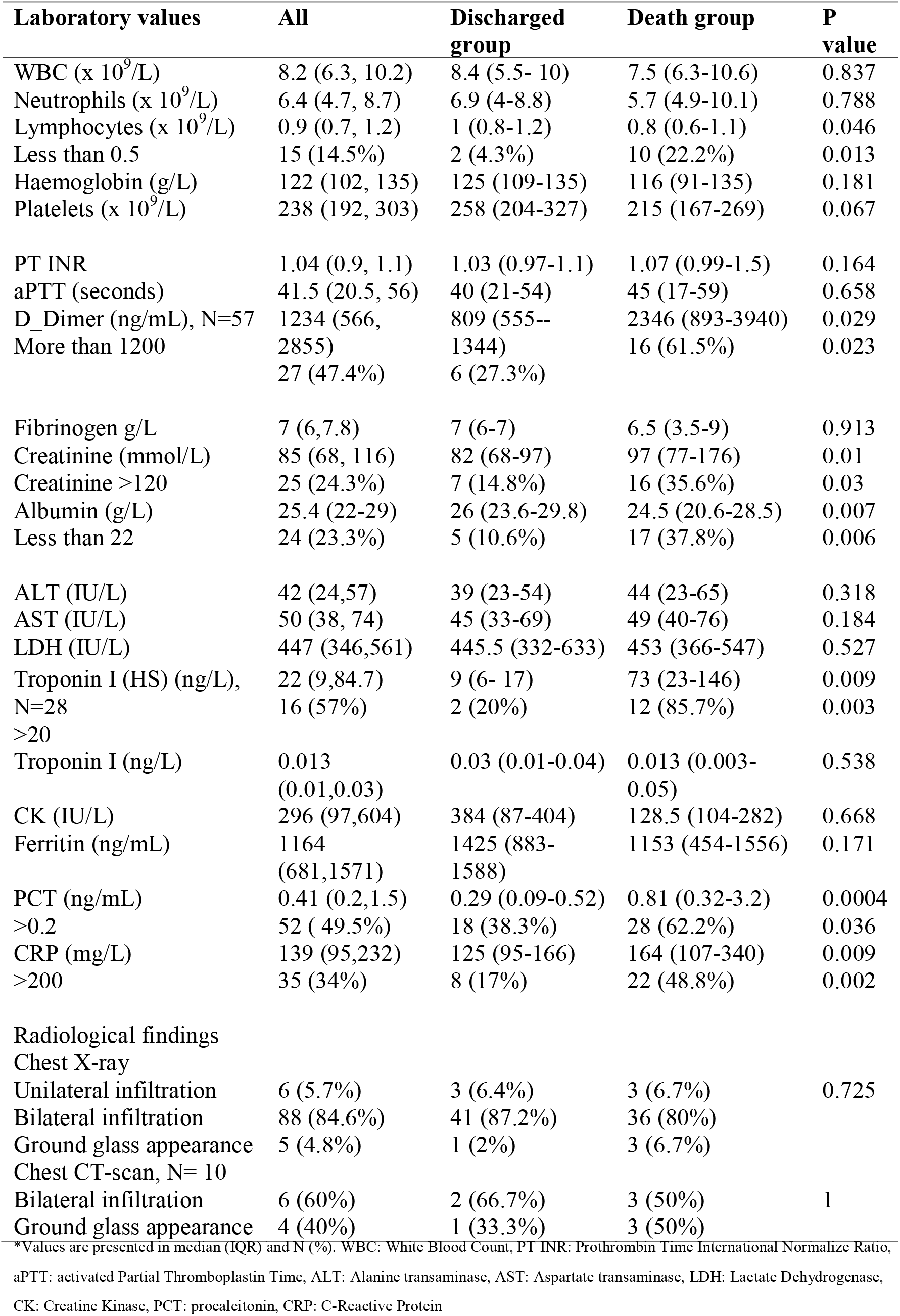
Laboratory and radiological findings on admission.

Majority of the patients (84.6% [87/103]) exhibited bilateral infiltrations on chest x-ray (Table 2). Only 10 patients had CT-scan results of the chest, with 60% of them showing bilateral infiltration and 40% showing ground glass appearance. There was no significant difference between the groups on the basis of the radiological findings.

### 3.3. Treatment and clinical course

Treatment and clinical course are shown in Table 3. Most patients (96%) were treated with antibiotics.Antiviral therapy (ritonavir-lopinavir) was used for 82.5% of the patients, while oseltamivir was used for 27% of the patients. Hydroxychloroquine was administered in 78% of the patients. Among the 103 patients, only 14.6% received systemic steroids. Majority of the patients (76.7%) were intubated and mechanically ventilated, and 50% of the patients wereprone positioned.A total of nine patients (8.7%) required ECMO, nine patients (8.7%) received inhaled nitric oxide (iNO), and three patients (2.9%) received convalescent plasma.

**Table 3:** Treatment and clinical course.

|  | All | Discharged | Death | P value |
| --- | --- | --- | --- | --- |
| Lopinavir -Ritonavir | 82 (79.6%) | 41 (87.2%) | 34 (75.5%) | 0.184 |
| Oseltamivir | 28 (27.2%) | 16 (34%) | 10(22.2%) | 0.251 |
| Hydroxychloroquine | 79 (76.7%) | 32 (68%) | 37 (82.2%) | 0.151 |
| Antibiotics | 99 (96%) | 45 (96%) | 45 (100%) | 0.495 |
| Inotropic support | 69 (71%) | 22 (50%) | 38 (86.4%) | <0.001 |
| Systemic steroid | 15 (14.6%) | 7 (14.9%) | 8 (17.8%) | 0.782 |
| Convalescent plasma transfusion | 3 (3%) | 0 | 3 (8%) | 0.092 |
| Maximum respiratory support |  |  |  |  |
| • High flow nasal canula | 22 (21.4%) | 17 (36.2%) | 0 |  |
| • Non-invasive ventilation | 2 (1.9%) | 2 (4.3%) | 0 |  |
| • Intubation | 79 (76.7%) | 28 (59.6%) | 45 (100%) | 0.001 |
| Prone positioning, N=95 | 51 (53.7%) | 22 (45.5%) | 23 (54.8%) | 0.518 |
| ECMO | 9 (8.7%) | 5 (10.6%) | 3(6.7%) | 0.714 |
| iNO | 9 (8.7%) | 4 (8.5%) | 5 (11%) | 0.737 |
| RRT | 21 (20.4%) | 3 (6.4%) | 17 (37.8%) | 0.001 |
| ARDS N= 96 |  |  |  |  |
| None | 11 (11.5%) | 11 (26%) | 0 |  |
| Mild | 12 (12.5%) | 5 (11.9%) | 6 (13.9%) | 0.002 |
| Moderate | 53 (55%) | 19 (45.2%) | 27 (62.8%) |  |
| Severe | 20 (21%) | 7 (16.7%) | 10 (23.3%) |  |
| Moderate-Severe ARDS | 73 (76%) | 26 (62%) | 37 (85%) | 0.014 |
| Secondary bacterial infection | 26 (25%) | 7 (14.8%) | 16 (35.5%) | 0.03 |
| Length of ICU stay, days | 11 (6-18.5) | 10 (6-15) | 13 (7-23) | 0.244 |

8Data are presented in N (%) and median (IQR). ECMO: extracorporeal membrane oxygenation, iNO: inhaled Nitric Oxide, RRT: Renal replacement therapy, ARDS: Acute Respiratory Distress Syndrome

Moderate/ severe ARDS was present in 76% of the patients, and 25% patients developed secondary bacterial infection. Inotropic support was used for 73% of the patients, and the proportion of deceased patients who required inotropic support was significantly higher than that in the discharged patients (86.4% versus 50%; P <0.001). Renal replacement therapy (RRT) was used in 20.4% of the patients and two of the patients who had pre-existing chronic kidney disease needed RRT. Moreover, among the deceased patients,intubation, RRT, moderate/severe ARDS,and secondary bacterial infection were observed more frequently than the discharged patients. Median time to discharge from ICU was 10 days (IQR:6-15), and median time to death was 13 days (IQR: 7-23).

### 3.4. Predictors of mortality

The multivariate logistic regression analysis showed thatthe risk factors associated with mortality were pre-existing hypertension (OR:3.2,95%CI: 1.2-8.9), moderate/severe ARDS (OR:3.4, 95%CI: 1.1-10.8), lymphocyte counts <0.5 × 10^9^/L (OR: 6.1, 95%CI: 1.2-29.8), albumin < 22 g/L (OR: 7.5, 95%CI: 2.1-26.2), procalcitonin (PCT)>0.2ng/mL (OR: 3.8, 95%CI: 1.3-7.8), D-Dimer >1200 ng/mL (OR: 5.1, 95%CI: 1.2-21.6), and the need for RRT(OR: 9.3, 95%CI: 2.4-36.2) (Table 4).

**Table 4:** Predictors to mortality.

| Predictor | OR (95%CI); P value |
| --- | --- |
| Age |  |
| Less than 40 | Reference |
| 41-50 | 5.5 (0.95 to 31.9);0.057 |
| 51-60 | 5.4 (0.95 to 31.2);0.058 |
| 61-70 | 3.5 (0.55 to 21.8);0.187 |
| More than 71 | 4.7 (0.51 to 44.2);0.174 |
| Comorbidities |  |
| 0 | Reference |
| 1 | 1.3 (0.41 to 4.1);0.676 |
| 2 | 2.9 (0.9 to 9.2);0.072 |
| **Pre-existing hypertension** | **3.2 (1.2 to 8.9); 0.02** |
| qSOFA > 2 points | 2.2 (0.95 to 5.2);0.065 |
| **Moderate to severe ARDS** | **3.4 (1.1 to 10.8);0.038** |
| Creatinine >120 mmol/L | 3.1 (0.92 to 9.9);0.069 |
| **Albumin <22 g/L** | **7.5 (2.1 to 26.2);0.002** |
| **Lymphocyte counts <0.5 x10<sup>9</sup>** | **6.1 (1.2 to 29.8);0.026** |
| **PCT>0.2 ng/mL** | **3.8 (1.3 to 7.8); 0.014** |
| CRP> 200 ng/ml | 2.3 (0.78 to 6.8);0.133 |
| **D- dimer >1200 ng/mL** | **5.1 (1.2 to 21.6); 0.026** |
| Sepsis | 2.7 (0.96 to 7.9);0.057 |
| Intubation | 2.1 (0.95 to 4.6);0.064 |
| Inotropic support | 3.5 (0.87 to 14.1);0.078 |
| **RRT** | **9.3 (2.4 to 36.2);0.001** |

## 4. Discussion

In this retrospective case series, we described the clinical characteristics and mortality-related predictors in adult patients with confirmed COVID-19 who were admitted to the critical care unit between 3^rd^March 2020 and 30^th^April 2020. The patient outcome was followed up to 20^th^ May 2020. The major outcome assessed here was patient mortality. The median age of our patients was 53 years. We evaluated the association between the patient’s demographic, laboratory, and clinical data and mortality. We found that, similar to other studies, there was no correlation between mortality and gender. However, unlike other studies, we did not find any significant correlation between advancing age and patient mortality. This finding could be attributed to the fact that most of the patients recruited were aged between 44 and 63 years, while other studies include higher proportion of individuals >65 years of age, which results in more significant association between age and mortality [7, 12].

Several studies have shown that presence of comorbidities, such as cardiovascular disease and secondary infections, contributes significantly to disease severity and mortality among COVID-19 patients [7,13,14]. In our study, we found that 37% of the patients who were admitted to the ICU exhibited presence of previous comorbidities, the most common being diabetes (39.2%) and hypertension (35%). Similarly, in the death group, most of the patients suffered from hypertension (51.1%) and diabetes (51.1%). This finding was consistent with those of previous studies [14]. Furthermore, our results also indicated hypertension to be an independent risk factor for mortality (adjusted OR: 3.2; 95%CI: 1.2-8.9). Moreover, compared to the discharge group, more than double the number of patients in the death group suffered from two or more comorbidities.

In several studies, the ICU mortality has been associated with organ dysfunction. qSOFA is a recently developed scoring system used to assess the organ dysfunction. Although qSOFA score does not directly evaluate patient mortality, previous studies have demonstrated the relationship between mortality and dysfunction [15]. In our study too, higher proportion of the patients in the death group showed higher qSOFA scores compared to the patients in the discharged group (35.6% vs. 22.2%, respectively); however, the difference was not statistically significant (P = 0.245).

Our results also indicated that the patient mortality was significantly associated with higher levels of (HS) Troponin, PCT, and CRP. Elevated (HS) troponin levels have previously been associated with acute coronary syndrome, reduced left ventricular function, and higher levels of IL-6 and TNF-alpha, which ultimately leads to mortality among the ICU patients [16]. PCT, precursor of calcitonin, is mainly released during bacterial sepsis and tissue injury. It has previously been recognized as an indicator of severity of organ failure and mortality among the ICU patients [17,18]. It is noteworthy that high PCT levels are associated with bacterial infection, which indicates that the death among the COVID-19 patients with elevated PCT might be due to secondary bacterial infections. This assumption was supported by our results that showed that compared to the discharge group, higher proportion of patients in the death group suffered from bacterial infections. CRP is a well-known plasma protein, which is involved in inflammatory reactions, and elevated CRP levels have been significantly associated with several cardiovascular disorders, organ failure, and mortality among the ICU patients [19,20].

Furthermore, lower levels of lymphocytes (lymphopenia) and albumin have also been associated with patient mortality. Within the death group, 37.8% and 22.2% of the patients exhibited albumin < 22 g/L and lymphocytes < 0.5 × 10^9^/L, which was significantly higher compared to the discharge group (10.6% and 4.3%, respectively) (P < 0.05). Our results corroborated with those of previous studies [21,22]. Furthermore, D-Dimer has been previously shown to a sensitive independent predictor of mortality among ICU patients [23], which was corroborated by our results. We observed that, compared to the discharge group, significantly higher proportion of patients in the death group exhibited D-Dimer > 1200 ng/mL (P = 0.023). However, a few studies have discouraged the use of D-Dimer quantification for mortality prediction in ICU patients [24].

With respect to the treatment approach adopted, we found that the requirement of inotropic support and intubation was significantly higher among the patients of the death group. In addition, RRT requirement was also found to be significantly associated with patient mortality, even after adjusting for the confounding factors (adjusted OR: 9.3; 95%CI: 2.4-36.2). It is worth mentioning that although high flow nasal cannula and non-invasive mechanical ventilation were only used in 21% and 2% of our patients respectively, none of them died, this could indicate a survival benefit for patients who receive non-invasive ventilation. However, it is very difficult to draw this conclusion at this point due to the nature of our study, and such result could be confounded.

There were several limitations of our study. Firstly, the sample size in our study was relatively small. Recruitment of a higher number of patients could have provided a clearer picture. Secondly, the patient data was collected retrospectively from their electronic records, which often had incomplete information. This could lead to information bias. Thirdly, there was no standard protocol for laboratory workup of the patients during ICU admission. This led to the absence of the results of some laboratory tests for some patients. Due to this, the laboratory-based data for some patients could not be included in the regression model, which might lead to discrepancy with respect to mortality predictors. Finally, at the end of the follow-up period, 11 patients were still in ICU, among which six were intubated, and one was on ECMO. This could again lead to bias in our final analysis as the mortality among these patients remained undefined. We propose for the future studies to recruit higher number of patients and to follow them up for a longer duration to further elucidate the factors that contribute to mortality among the COVID-19 patients.

## 5. Conclusion

Our observational study involving critically ill patients with confirmed COVID-19 showed no significant differences in mortality based on age, gender, or nationality. Several risk factors were identified to predict mortality. Based on our results, we established a set of predictive factors “HARD PAL,” which can be used as a tool to predict mortality in critically ill COVID-19 patients. More multicentric large studies are needed to validate the significance of these predictors with respect to patient mortality.

|  |  |
| --- | --- |
| **H** | Pre-existing **hypertension** |
| **A** | Severe **ARDS** (PO<sub>2</sub>/FiO<sub>2</sub> is less than 100) |
| **R** | **Renal** replacement therapy (RRT) |
| **D** | **D-dimer** more is than 1200 ng/ml |
| **P** | **Procalcitonin** level is more than 0.2 ng/ml |
| **A** | Serum **albumin** is less than 22 g/L |
| **L** | **Lymphocytic** count is less than 0.5. 10<sup>9</sup>/L |

## Data Availability

Data are available

## Abbreviations

(ICU): Intensive care unit
(RT-PCR): Real-time reversetranscriptasepolymerasechainreaction
(RRT): Renal replacement therapy
(ECMO): Extracorporeal membrane oxygenation
(ARDS): Acute respiratory distress syndrome
(qSOFA): Quick Sequential Failure Assessment
(ACEI): Angiotensin converting enzyme inhibitors
(PCT): Procalcitonin
(CRP): C-reactive protein

## Acknowledgment

We thank Dr. Abdulrahman Alrefai, Dr. Ali Bander and Dr. Abdulrahman Alfares who designed the record sheet for data collection.

